# Treadmill Exercise Stress Echocardiography Exposes Impaired Left Ventricular Function in Patients Recovering from Hospitalization with COVID-19 Without Overt Myocarditis Versus Historical Controls

**DOI:** 10.1101/2024.02.01.24302037

**Authors:** Robert E Goldstein, Edward A. Hulten, Thomas B. Arnold, Victoria M. Thomas, Andrew Heroy, Erika N. Walker, Keiko Fox, Hyun Lee, Joya Libbus, Bethelhem Markos, Maureen N. Hood, Travis E. Harrell, Mark C. Haigney

## Abstract

**Background:** Usual clinical testing rarely reveals cardiac abnormalities persisting after hospitalization for COVID-19. Such testing may overlook residual changes responsible for increased adverse cardiac events post-discharge.

**Methods:** To further elucidate long-term status, we performed exercise stress echocardiography (ESE) in 15 patients age 30-63 without myocarditis 3 to 31 months after hospital discharge. We compared patient outcomes to published data in healthy comparisons (HC) exercising according to the same protocol.

**Results:** Patients’ treadmill exercise (Bruce protocol), averaging 8.2 min, was halted by dyspnea or fatigue. Pre-stress baselines in recovering patients (RP) matched HC except for higher heart rate: mean 81 bpm for RP and 63 for HC (p<0.0001). At peak stress, RP had significantly lower mean left ventricular (LV) ejection fraction (67% vs 73%, p<0.0017) and higher peak early mitral inflow velocity/early mitral annular velocity (E/e’, 9.1 vs 6.6, p<0.006) compared with HC performing equal exercise (8.5 min). Thus, when stressed, patients without known cardiac impairment showed modest but consistently diminished systolic contractile function and diastolic LV compliance during recovery vs HC. Peak HR during stress was significantly elevated in RP vs HC; peak SBP also trended higher. Average pulmonary artery systolic pressures among RP remained normal.

**Conclusions:** Our measurements during ESE uniquely identified residual abnormality in cardiac contractile function not evident in the unstressed condition. This finding exposes a previously-unrecognized residual influence of COVID-19, possibly related to underlying autonomic dysfunction, microvascular disease, or diffuse interstitial changes after subclinical myocarditis; it may have long-term implications for clinical management and later prognosis.

**CLINICAL PERSPECTIVE:** New Findings (relative to a historical comparison group)

- Symptom-limited treadmill exercise 3-31 months after hospitalization with COVID-19 without overt myocarditis elicited a lesser rise in left ventricular ejection fraction than seen in similar subjects with no exposure to COVID-19.
- The same symptom-limited exercise in these patients revealed evidence of diminished left ventricular diastolic function relative to subjects with no exposure to COVID-19.
- These distinctive differences in left ventricular function were observed although overall exercise capacity was the same as in the uninfected comparison group.

Clinical Implications

- Prior hospitalization with COVID-19 even in the absence of overt myocarditis was often associated with a modest but consistent decrement in left ventricular systolic contraction and diastolic relaxation; these functional abnormalities were evident after peak treadmill exercise stress despite lack of distinctive difference in contractile parameters at rest.
- Patients recovering after hospitalization with COVID-19 may benefit from sustained observation of their cardiovascular status and adjustment of their exercise requirements appropriate to individual cardiovascular capabilities.
- Treadmill stress testing with echocardiography uniquely identifies potentially important differences in the cardiovascular function of patients recovering after hospitalization with COVID-19.

## INTRODUCTION

COVID-19 can adversely affect the heart and circulation, particularly in hospitalized patients (1). Early experience demonstrated that severe acute COVID-19 infection can lead to heart failure and arrhythmia (2). Cardiovascular abnormalities may persist into the recovery period and contribute to disability lasting many months. Recent studies have shown substantially increased occurrence of multiple different adverse clinical cardiovascular consequences in the year after hospitalization for COVID-19 compared with COVID-infected but non-hospitalized patients and compared with uninfected patients (3). While lingering direct viral or immune actions may play a role, the reasons for these persistent cardiovascular problems and their long-term implications remain unclear.

Routine testing of blood troponin, brain natriuretic peptide, and C-reactive protein shows no abnormalities during recovery in nearly all patients hospitalized with COVID-19 without overt myocarditis during or after their hospital stay (4–5). The same lack of abnormality is also observed when these patients have electrocardiograms, echocardiograms, or magnetic resonance images (6–8). However, prior investigations have not assessed the performance of the heart and circulation during maximal exercise-induced stress, which may bring out abnormalities that are inapparent in the absence of such stress. Treadmill performance limited by cardiovascular symptoms such as fatigue or dyspnea is likely to elicit maximal cardiac function, as evidenced by substantially increased left ventricular ejection fraction (LVEF) and stable diastolic performance parameters (9). Sensitive detection of cardiovascular abnormalities after hospitalization with COVID-19 may have value in identifying those at risk for later complications. In addition, assessment of exercise stress performance may have benefit in choosing proper activities for recovering patients. Accordingly, we evaluated treadmill echocardiographic performance at least 6 weeks after hospital discharge using standard clinical methods in patients without evidence of myocarditis during or after hospitalization for COVID-19.

## METHODS

### Hypotheses

Utilizing stress echocardiography (ESE), we assessed whether cardiac contractile performance at symptom-limited maximum effort among recovering patients (RP) after hospitalization with COVID-19 would differ from published data (9) for similarly-elicited ESE results in healthy comparison (HC) subjects with no exposure to COVID-19. Our pre-specified primary hypothesis was that RP and HC would show no statistically significant difference in LVEF soon after maximum stress. Our secondary hypothesis was that measurements made during the same maximal stress assessment would demonstrate no statistically significant difference in peak early mitral inflow velocity (E) over the early diastolic mitral annular velocity (e’), E/e’, between RP and HC.

### Patient recruitment

Advertisements were posted in appropriate outpatient clinics at Walter Reed National Military Medical Center, and referrals were solicited from staff physicians who managed care for patients admitted to hospital with COVID-19. All patients eligible for military health care and age <65 years without prior cardiovascular diagnosis other than essential hypertension were eligible for study enrollment >6 weeks post-discharge from hospital after admission with a positive PCR test for COVID-19. Each study enrollee had an interview, an ECG, and serum troponin, brain natriuretic peptide, and d-dimer levels at enrollment.

Prior to the COVID-19 pandemic, 31 healthy comparison (HC) subjects were assessed according the same ESE protocol used in the present study (9). Their performance is taken as comparison data for our patients studied after hospitalization with COVID-19.

### Stress echo methods

15 of 20 enrolled patients were deemed appropriate for ESE based on 1) lack of evidence of myocarditis on current testing (ECG and troponin levels) or prior in-hospital results (ECG, troponin, or MRI) indicative of myocarditis and 2) patient-observed ability to walk a mile or climb 3 flights at a normal pace without pause at the time of study enrollment. All ESE testing was supervised and interpreted by experienced, echo board certified cardiologists (EH and TH) and other clinical personnel according to a pre-specified protocol used for routine clinical ESE that included baseline blood pressure, SpO2, ECG, and cardiac ultrasound measurements. Patients then exercised on a motor-driven treadmill according to standard Bruce protocol with continuous ECG monitoring and cuff measurement of blood pressure every 3 minutes and just before stopping.

Exercise was continued until halted by symptoms judged to preclude further effort. The final echocardiogram was recorded within 2 minutes of stopping exercise.

Imaging and Doppler echocardiograms were recorded using a single commercially-available unit. LVEF was calculated from the 2-dimensional echocardiography data using the American Society of Echocardiography recommended technique of biplane method of disks summation (modified Simpson’s rule). For diastolic evaluation using an apical window, the pulsed Doppler sample volume was placed at the mitral valve tips, and 5 to 10 cardiac cycles were recorded and averaged. The mitral inflow velocities were recorded, and the peak velocity of early filling (E) was derived. Special pulse-wave Doppler mode was used to assess tissue velocities. For measurements of mitral annular velocities (e’), the filter was set to exclude high-frequency signals and a Nyquist limit adjusted to 15-20 cm/s. Gain was reduced to obtain a clear tissue signal with minimal background noise. From the apical 4-chamber view, a 2- to 5-mm sample volume was placed at the septal corner of the mitral annulus. The E/e’ ratio was calculated from paired mean values of E and e’. Maximal tricuspid regurgitation (TR) velocity was evaluated in apical and subcostal views; for each patient, maximal TR velocity was averaged over a single respiratory cycle. Right atrial pressure (RAP) was estimated using inferior vena cava size and inspiratory collapse in subcostal view (10–11). Pulmonary artery systolic pressure (PASP) was calculated as 4(TR_vmax_)^2^ + RAP as per usual standard practice. The measurements just described were obtained in the same sequence before and after exercise.

### Statistical methods

In this study, we aimed to investigate the potential associations between various clinical parameters and outcomes in patients hospitalized with COVID-19. After accounting for missing data, the resulting sample size was deemed insufficient to support multivariate analysis. Therefore, we opted to perform univariate analyses, which, despite their limitations, allowed us to explore individual relationships between the variables and the primary outcome. Univariate correlations and significance between chosen outcomes were calculated. Mean values are shown ± standard deviation. After checking for the normality of comparator variables within the RP sample (LVEF at rest, stress LVEF, E/e’ at rest, and stress E/e’) using the Shapiro-Wilk test, corresponding mean values for RP and HC were compared using the unpaired Student’s t-test, which was deemed valid based on generally similar sample sizes and variances for the given comparator variables. Values for all statistical tests are deemed statistically significant if p<0.05. Future studies with a larger and more diverse sample are required to fully investigate the potential interplay of factors involved in this condition. The limited sample size prevented the application of linear modeling.

The protocol for this study (WRNMMC-2020-0319) received final approval from the Walter Reed Investigational Review Board on Dec 1, 2020. Patient enrollment occurred May 10, 2021-June 30, 2022.

## RESULTS

### Patients (Table 1)

Recovering patients (RP) after COVID-19 were age 49±11 years; age range was 30-63 years. Nine (60%) were women and six were men. Seven (47%) were active-duty servicemembers. Body mass index averaged 33.7±6.2 kg/m^2^; 8 patients were >30 kg/m^2^. (Table 1). Eight (53%) were on antihypertensive medications, including 3 on beta blocking drugs (dose held the morning of ESE). One patient, a woman in her 40’s, took medications to control diabetes. This patient had no known prior cardiovascular disorder besides mild, uncomplicated essential hypertension.

**Table 1.**
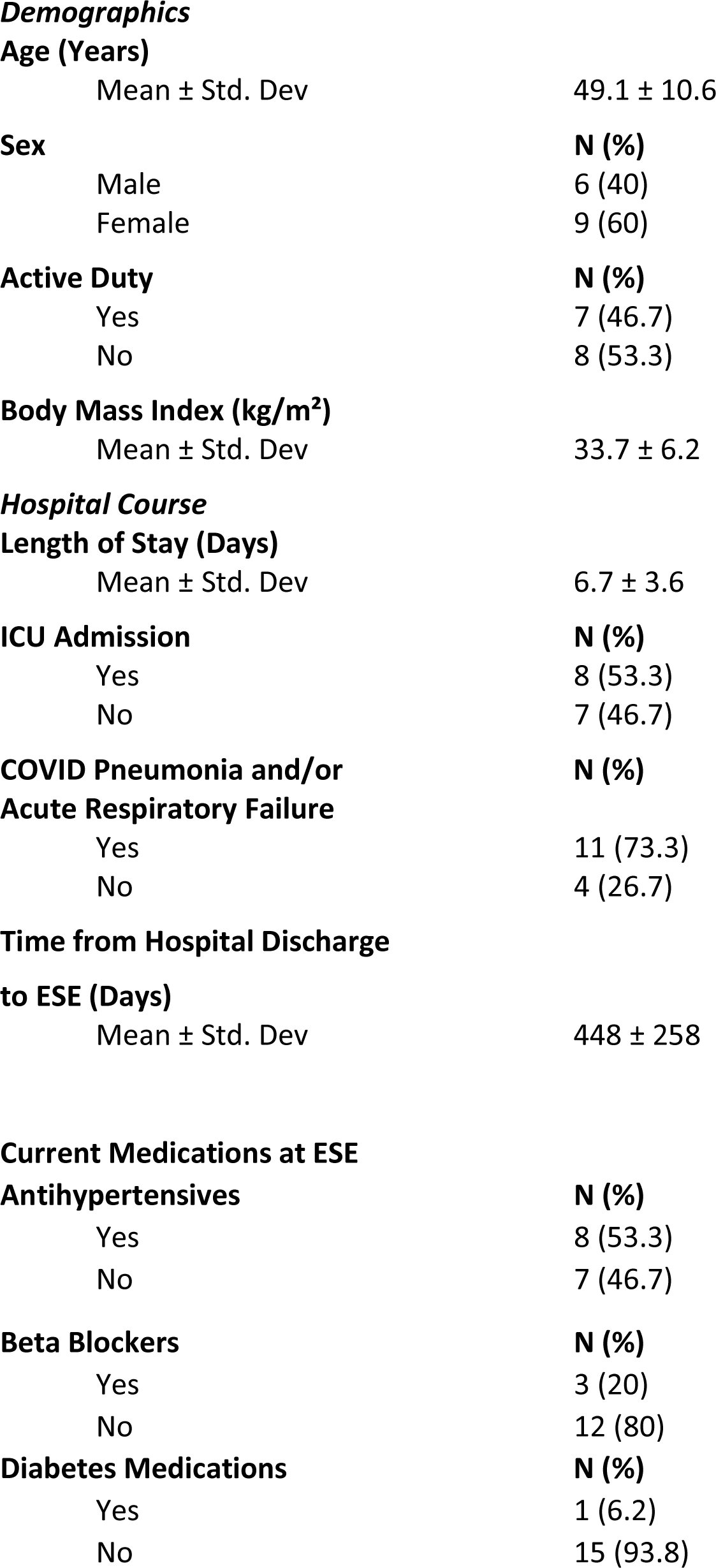
Study Subjects Having Exercise Stress Echocardiography (n=15)

RP were admitted to hospital between April 2020 and January 2022; each had a positive PCR test for COVID-19 when hospitalized. Twelve (80%) had no prior covid-19 vaccination and none had known prior infection with COVID-19. Eight (53%) were treated in the intensive care unit. Total hospital stay averaged 6.7±3.6 days. Principal discharge diagnosis was COVID-19 pneumonia in 10 (67%) with accompanying acute hypoxic respiratory insufficiency in 4. Other individual patients were admitted for pericarditis without myocarditis, branch-vessel pulmonary embolism, neurological symptoms, pancreatitis and severe emesis.

Prior to enrollment, all patients provided written informed consent. At study enrollment, all 15 having ESE had a normal electrocardiogram and normal values for serum high-sensitivity troponin T, brain natriuretic peptide, and d-dimer.

Historically-derived healthy comparison (HC) subjects were age 59±14 years and 18 (58%) were women. None had hypertension and none took medications.

### Exercise Stress Test (ESE)

ESE testing was performed 448±258 days after hospital discharge (range 97-949 days). All patients wore face masks at rest and during exercise, in keeping with hospital safety regulations. At outset, all 15 patients had normal (>95%) SpO2.

Patients walked on a treadmill at increasing speeds and grades following the standard Bruce protocol until halted by symptoms. These symptoms included limiting fatigue and dyspnea in each; some subjects had accompanying leg pain or pleuritic-type chest pain. None had ischemic-type chest pain. All patients remained in sinus rhythm during exercise and recovery; none had associated abnormal ST-T wave changes. All study patients had post-exercise echocardiograms recorded within two minutes of halting exercise. HC performed treadmill exercise according to the same Bruce protocol and were limited by fatigue or dyspnea. Echo methods for HC were the same as those used for RP (9).

At supine rest before exercise, RP had hemodynamic parameters the same as HC, except for a higher mean heart rate (Table 2, upper panel and Figure 1). Specifically, left ventricular ejection fraction (LVEF) and E/e’ (peak early mitral inflow velocity [E] over the early diastolic mitral annular velocity [e’]), were not different from HC. PASP, estimated at rest in 8 patients, was normal (<36 mmHg)(11) in each individual (group mean 25 ±6 mmHg). Recovering patients’ regional wall motion, assessed in each of the 15, was normal at rest and after treadmill stress and no chamber enlargement was observed.

**Figure 1.**
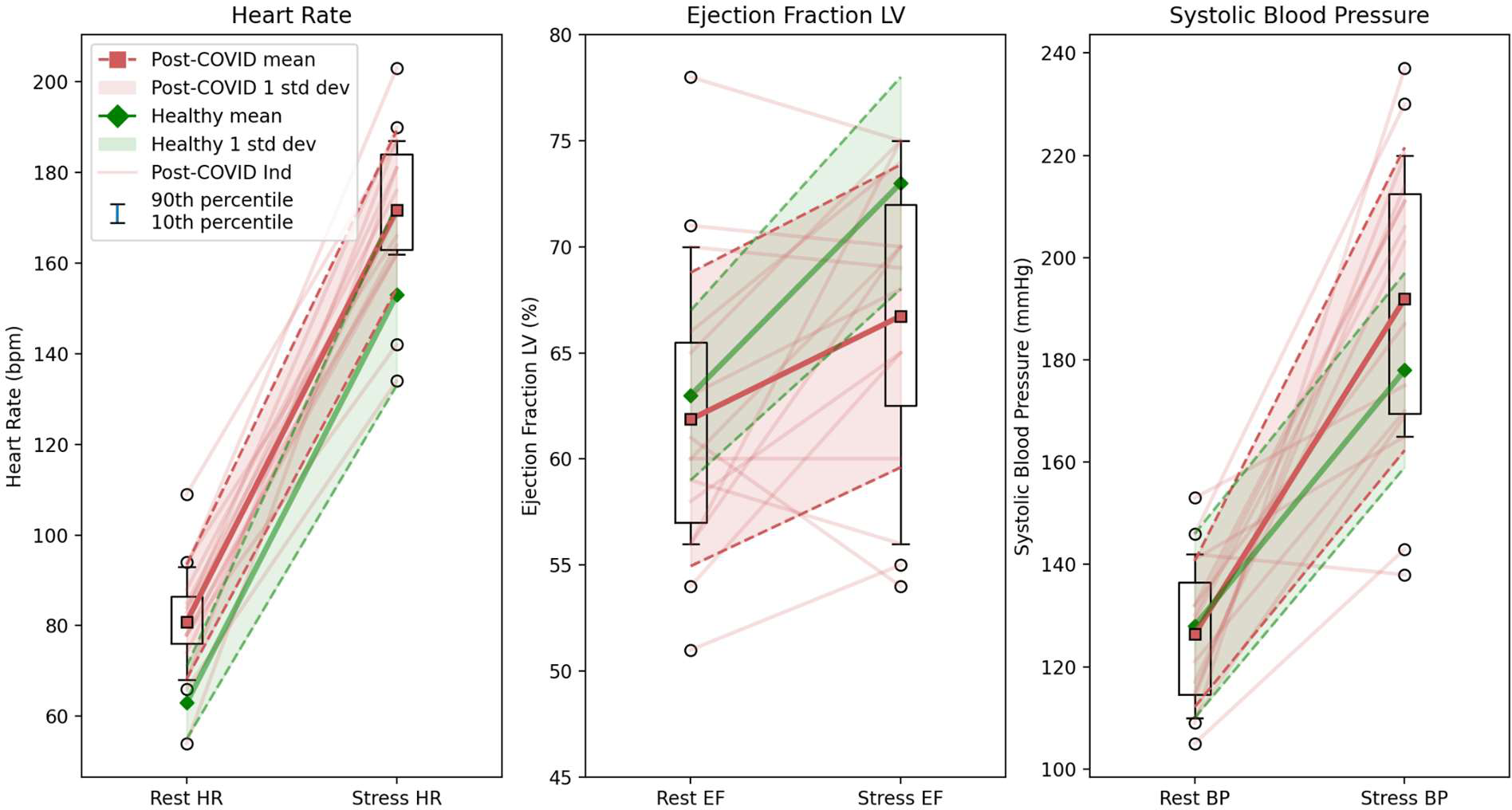
**Legend:** In each panel, values at rest pre-stress (left) are connected to corresponding values at peak symptom-limited treadmill stress (right). Figure 1 depicts heart rate (left panel), left ventricular ejection fraction (LVEF, middle panel), and systolic blood pressure (BP, right panel). Data for recovering patients (RP) are shown in red, and data for healthy comparison (HC) subjects in green. Means are represented by squares and heavy lines; one standard deviation is shown as shaded areas bounded by dashed lines. For RP data, “box and whisker” plots denote the 25^th^ and 75^th^ percentiles (bottom and top of boxes) plus 10^th^ and 90^th^ percentiles (whiskers). “Fliers” outside those whiskers are shown as circles. Individual patient performance is indicated by fine lines. Mean stress LVEF for RP is significantly below stress LVEF for HC. Heart rate is also significantly higher at rest and peak stress. Exercise BP tends higher in RP but the difference is not statistically different.

**Table 2.**
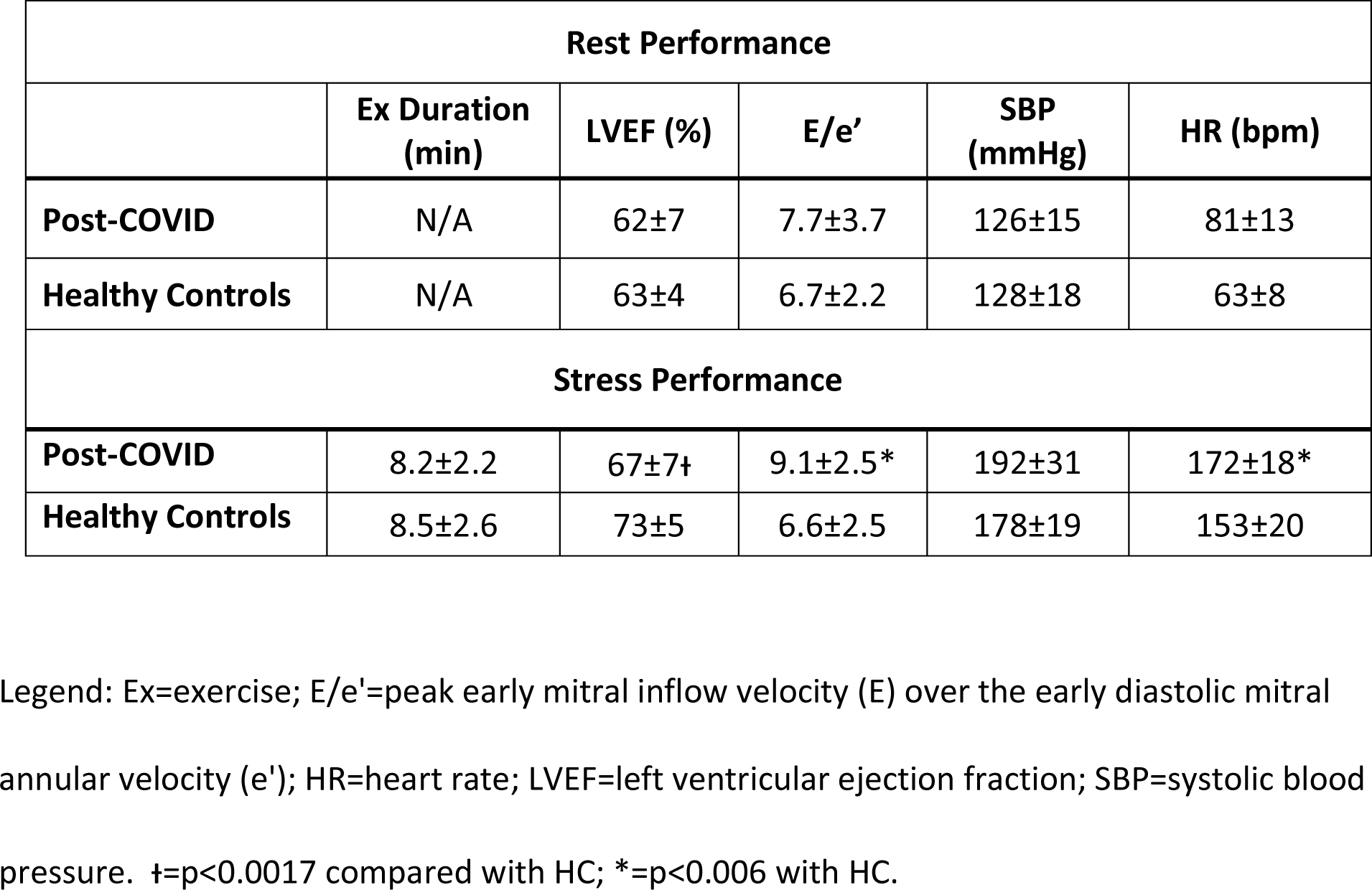
Cardiac Parameters at Pre-Stress Rest and Peak Treadmill Exercise.

| **Rest Performance** |  |  |  |  |  |
| --- | --- | --- | --- | --- | --- |
|  | **Ex Duration (min)** | **LVEF (%)** | **E/e'** | **SBP (mmHg)** | **HR (bpm)** |
| **Post-COVID** | N/A | 62±7 | 7.7±3.7 | 126±15 | 81±13 |
| **Healthy Controls** | N/A | 63±4 | 6.7±2.2 | 128±18 | 63±8 |
| **Stress Performance** |  |  |  |  |  |
| **Post-COVID** | 8.2±2.2 | 67±7† | 9.1±2.5* | 192±31 | 172±18* |
| **Healthy Controls** | 8.5±2.6 | 73±5 | 6.6±2.5 | 178±19 | 153±20 |

Legend: Ex=exercise; E/e'=peak early mitral inflow velocity (E) over the early diastolic mitral annular velocity (e'); HR=heart rate; LVEF=left ventricular ejection fraction; SBP=systolic blood pressure. †=p<0.0017 compared with HC; \*=p<0.006 with HC.

Recovering patients performed according to the same exercise protocol as HC and stopped due to limiting fatigue or dyspnea after mean 8.2 ± 2.2 minutes, nearly the same as the symptomatic stopping time for HC (Table 2, lower panel). By contrast, mean final LVEF for RP was significantly lower (66.7 ±7.4% RP vs. 73 ±5% HC, p=0.0014) and mean E/e’ (n=14) significantly higher (9.1±2.5 RP vs. 6.6±2.5 HC, p=0.0031) (Figure 1, middle panel). In addition, mean peak heart rates were significantly higher (172±18 bpm RP vs 153±20 HC, p=0.0032). Systolic blood pressure values at end-exercise tended higher in RP compared with HC, but were not consistently so (p>0.05). Among study subjects, Average PASP for RP (n=13) at end-exercise was 31±9 mmHg. In 6 RP with paired estimates, mean rise in PASP was 5.7±1.7 mmHg (p=0.023).

Correlation coefficients and their associated p-values were calculated for RP (Table 3). This correlation analysis indicated no association of stress LVEF in study patients with BMI, prior use of antihypertensive medication, age, gender, or systolic blood pressure. The correlation analysis also indicated no association of stress E/e’ with BMI, age, gender, or systolic blood pressure.

**Table 3.** Correlation Matrix of Selected Patient Variables.

|  |  |  |  |  |  |  |  |
| --- | --- | --- | --- | --- | --- | --- | --- |
| **Age** | 1 |  |  |  |  |  |  |
| **Gender** | .018 | 1 |  |  |  |  |  |
| **BMI** | 0.277 | -0.134 | 1 |  |  |  |  |
| **Anti-HTN Med** | **0.600\*** | 0.043 | **0.573\*** | 1 |  |  |  |
| **Stress Systolic BP** | -0.109 | -0.013 | 0.031 | -0.124 | 1 |  |  |
| **Stress LVEF** | 0.137 | -0.382 | 0.043 | -0.247 | -0.101 | 1 |  |
| **Stress E/e'** | 0.227 | -0.09 | 0.219 | 0.32 | -0.22 | -0.19 | 1 |
|  | **Age** | **Gender** | **BMI** | **Anti-HTN Med** | **Stress Systolic BP** | **Stress LVEF** | **Stress E/e'** |

\*: Significant correlation (p<0.05)
**Legend:** Correlation matrix assesses the linkage of 7 clinical characteristics and hemodynamic values for the 15 recovering patients. Numerical values ranging from 1 to -1 indicate the strength of the paired linkage between horizontally and vertically arrayed features (identity=1). Antihypertensive therapy was significantly correlated with age and body mass index. However, Stress LVEF and E/e' were not correlated with any of the clinical parameters. Abbreviations: Anti-HTN Med=antihypertensive medications; BMI=body mass index; BP=blood pressure; E/e'=peak early mitral inflow velocity divided by early mitral annular velocity; LVEF=left ventricular ejection fraction.

### Follow-up

All study patients had 6-monthly structured telephone interviews to assess functional capacity and clinical occurrences. Each of the 15 was followed for at least a year after study enrollment. Elicited information covered 20.1 patient-years. RP had no deaths, myocardial infarctions, or strokes and no symptoms suggesting heart failure or sustained arrhythmia. Two of the 15 were hospitalized, one patient twice for pneumonia. At completion of follow-up for the 15 (averaging 1.3 years), seven were deemed to function in New York Heart Association Class 1 and eight in NYHA Class 2.

## DISCUSSION

Our 15 middle-aged study patients recovering from severe COVID-19 without overt myocarditis achieved the same duration of treadmill exercise as comparable uninfected individuals. However, their cardiovascular parameters at this same peak exercise were distinctly different: recovering patients had mildly but significantly diminished immediate post-exercise left ventricular performance, evident in both systolic contraction—lesser LVEF—and diastolic relaxation—increased E/e’. In addition, recovering patients had heart rates at peak exercise significantly higher than normal comparison. Patients’ systolic blood pressure tended to be higher, but this difference was not consistent.

Subnormal left ventricular performance at peak treadmill exercise was the only sign of cardiovascular dysfunction evident in our recovering patients. Echo parameters at rest, troponin levels, and the electrocardiogram showed no distinctive abnormality. Notably, our findings uniquely identified consistent subtle abnormalities of contractile function after severe COVID-19 even in patients with no evidence of myocarditis on routinely-surveyed parameters. These functional abnormalities seen only during treadmill stress suggest exercise testing is appropriate in the months after hospitalization for severe COVID-19 to identify those needing longer recovery before returning to physically demanding activity. Such testing may also aid in early identification of post-hospital patients at greatest risk of late-appearing COVID-related cardiac problems (3).

Our data permit a tentative assessment of possible causes for the observed abnormalities in cardiovascular function. While some of our patients were overweight or on treatment for hypertension, univariate correlation analysis indicated that neither these conditions nor age were strongly linked to the abnormalities observed in left ventricular function. Heart rates are often higher in patients symptomatic after COVID-19 at supine and upright rest (12), an effect that may be mediated in part by deconditioning. Blood pressure may also be increased late after COVID-19 (13). Our data may reflect greater adrenergic drive and lesser vagal tone in our patients. Such effects may figure in our observed augmentation of heart rate and blood pressure, possibly enhanced because our patients were wearing masks during exercise. However, mask-related enhancement of heart rate and blood pressure with exercise appears minor (14). Neither deconditioning nor increased adrenergic tone explains the changes we noted in systolic and diastolic left ventricular performance immediately after symptom-limited exertion: findings indicate maximal LVEF is not affected by training during maximal, symptom-limited treadmill exercise (15–16). Alternatively, the rises we observed in heart rate and blood pressure at maximal exercise may have been autonomically-mediated responses to exercise-induced myocardial dysfunction and resultant insufficient perfusion of exercising skeletal muscles. Overall, while not derived from a large sample size these interesting results add support to most recent American Society of Echocardiography recommendations for comprehensive exercise stress echocardiography involving not only systolic evaluation of regional wall motion and ejection fraction but also diastolic indices and pulmonary pressure at rest and with exertion (17).

Residual post-COVID pulmonary vascular dysfunction might interfere with normal left performance by inducing right ventricular overload. This is unlikely since our patients had normal pulmonary artery pressures at rest and immediately after maximal exercise. Normal peak pulmonary artery pressures also mitigate against exercise-induced hypoxia from residual lung abnormalities or mask-wearing. All our patients had normal SpO2 values at rest before stress; and other investigators found that mask-wearing causes only mild reductions in arterial oxygen content during exercise (14).

Myocardial ischemia might be a causal factor explaining our finding of impaired left ventricular performance during exercise following severe COVID. Coronary endothelial dysfunction (18–19) or diffuse interstitial changes in the myocardium after subclinical myocarditis may lead to insufficient rise in coronary blood flow during maximal exertion. Alternatively, persistent COVID-19 interference with oxidative metabolism of cardiomyocytes might limit high-energy phosphate synthesis in recovering patients (20–21), particularly during periods of high metabolic demand such as exercise. Neither the 12-lead electrocardiograms monitoring our patients during exercise nor the patients’ symptoms were indicative of myocardial ischemia. However, overt ischemic manifestations may not be apparent if the myocardial perfusion defect is modest or if impairment of oxidative metabolism in myocytes is mild.

## Limitations

Practical considerations related to the COVID-19 pandemic limited the number of recovering patients in this study and precluded contemporary controls. These findings are therefore not definitive and should be interpreted with caution, as analyses were conducted with a small sample size. Nevertheless, statistically significant differences emerged even with a limited number of study participants. Our study population may have experienced uniquely intense COVID-19 infection at hospitalization due to pandemic conditions and, for 80% of our patients, lack of prior vaccination. While the worst features of the COVID-19 pandemic have abated, the abnormality of cardiac function at peak exercise seen in our patients represents a previously-unrecognized aspect of COVID-19 infection, one that may have significant implications for select patients with ongoing COVID-19 vulnerability and for patients encountering future Coronavirus infections.

The comparison subjects taken as a comparison group were demographically close to our recovering patients. Their similar values for symptom-limited exercise duration and similar echocardiographic findings at rest favor the comparability of groups and confirm that the same ESE techniques were employed for the current studies and for the historical comparisons.

## Data Availability

There is no restriction regarding the availability of all data referred to in the manuscript.

## ABBREVIATIONS

E/e’: early mitral inflow velocity/early diastolic mitral annular velocity
ESE: exercise stress echocardiography
EF: ejection fraction
HC: healthy comparison subjects
HR: heart rate
LV: left ventricular
PASP: pulmonary artery systolic pressure
RAP: right atrial pressure
RP: recovering patients
TR: tricuspid regurgitation

## ACKNOWLEDGMENTS

The authors are grateful for the skilled input of Walter Reed echocardiographers, Ms. Hedda Richards and Ms. Teresa Collins.

